# Pharmacological and Non-Pharmacological Interventions to Enhance Sleep in Mild Cognitive Impairment and Mild Alzheimer’s Disease: A Systematic Review

**DOI:** 10.1101/2020.07.27.20162578

**Authors:** Jonathan Blackman, Marta Swirski, James Clynes, Sam Harding, Yue Leng, Elizabeth Coulthard

## Abstract

Suboptimal sleep causes cognitive decline and probably accelerates Alzheimer’s Disease (AD) progression. Several sleep interventions have been tested in established AD dementia cases. However early intervention is needed in the course of AD at Mild Cognitive Impairment (MCI) or mild dementia stages to help prevent decline and maintain good quality of life. This systematic review aims to summarize evidence on sleep interventions in MCI and mild AD dementia.

Seven databases were systematically searched for interventional studies where ≥75% of participants met diagnostic criteria for MCI/mild AD dementia, with a control group and validated sleep outcome measures. Studies with a majority of participants diagnosed with Moderate to Severe AD were excluded.

20164 references were returned after duplication removal. 284 full papers were reviewed with 12 retained. Included papers reported 11 separate studies, total sample (n=602), mean age 76.3 years. Nine interventions were represented: Cognitive Behavioural Therapy – Insomnia (CBT-I), A Multi-Component Group Based Therapy, Phase Locked Loop Acoustic Stimulation, Melatonin, Donepezil, Galantamine, Rivastigmine, Tetrahydroaminoacridine and Continuous Positive Airway Pressure (CPAP). Psychotherapeutic approaches utilising adapted CBT-I achieved statistically significant improvements in the Pittsburgh Sleep Quality Index with one study reporting co-existent improved actigraphy variables. Melatonin significantly reduced sleep latency and sleep to wakefulness transitions in a small sample. CPAP demonstrated efficacy in participants with Obstructive Sleep Apnoea. Evidence to support other interventions was limited.

There is a paucity of evidence for sleep interventions in MCI and mild AD highlighting a pressing need for high quality experimental studies exploring alternative sleep interventions.

## INTRODUCTION

The association between Alzheimer’s Disease [AD] and sleep disturbance is well established (Mander, 2013; Tranah et al., 2007). The traditional view has been that AD causes sleep impairment and the extent of symptomatic sleep disturbance correlates with the severity of dementia (Benca et al., 1992; Montplaisir et al., 1995; Pat-Horenczyk et al., 1998; Prinz et al., 1982; Weldemichael & Grossberg, 2010). Circadian rhythm disorders also contribute to sleep disturbance and worsen with age and AD, possibly related to alterations to the Suprachiasmatic Nucleus secretion of melatonin (Swaab et al. 1985).

A recent prominent theory postulates a bidirectional relationship between poor sleep and AD (Mander et al., 2016) - as well as AD causing sleep disturbance, sleep disturbance may lead directly to pathological accumulation of proteins that cause neurodegeneration (Fultz et al., 2019; Hahn et al., 2014). Disruption of sleep occurs early in AD (Ju et al., 2013; Lim et al., 2013; Sprecher et al., 2017), even before symptoms arise and AD pathology progresses for 1-2 decades before a full dementia diagnosis giving plenty of time for factors such as sleep disturbance to affect the rate of pathological progression. Regardless of whether sleep disturbance arises directly from accumulation of AD pathology or through an independent problem such as sleep apnoea (Ancoli-Israel, 2000; Bliwise, 1999; Van Cauter et al., 2000), poor sleep might be a significant contributory factor in the eventual onset and progression of AD.

Up to 60% of patients in memory clinics have sleep disorders (LittleJohn et al., 2014). Sleep deprivation brings a range of other metabolic and cardiovascular impairments that mean improving sleep is likely to be broadly beneficial in the memory clinic population over and above the potential to modify progression of AD. However, there are no standard sleep treatment protocols within memory clinics and other dementia services.

The optimal opportunity to reverse or halt cognitive decline caused by AD pathology, thus maintaining quality of life, will be early in the course of disease – at the Mild Cognitive Impairment or mild AD stage. Early diagnosis utilising molecular biomarkers to accurately pinpoint the cause of cognitive symptoms is becoming more common with early / prodromal AD the most frequently identified neurodegenerative cause. However, the focus of sleep intervention reviews to date have largely been at the stage of established AD dementia (Mitolo et al., 2018; O’Caoimh et al., 2019) when pathology may be too advanced to change.

There are multiple theoretical targets for sleep enhancement in AD (summarised in **Figure 1**). Much of the previous literature has focussed on pharmacological interventions (McCleery et al., 2016). Notable disadvantages of this approach, particularly in older adults, include the potential for side effects, interactions and polypharmacy (Maher et al., 2014). Hence, interest in non-pharmacological sleep-modifiers is intensifying with recent technological advances permitting exploration of novel approaches such as closed loop stimulation of slow wave sleep through sound or electrical brain stimulation (Ngo et al., 2013) and glasses to deliver Bright Light Therapy (Sekiguchi et al., 2017). Lifestyle interventions are often cost effective and non-toxic and have the potential to enhance sleep in early stages of AD, but have not previously been the focus of reviews in mild AD and MCI. We include all potential types of intervention, including lifestyle interventions, in the searches for this review.

**Figure 1.**
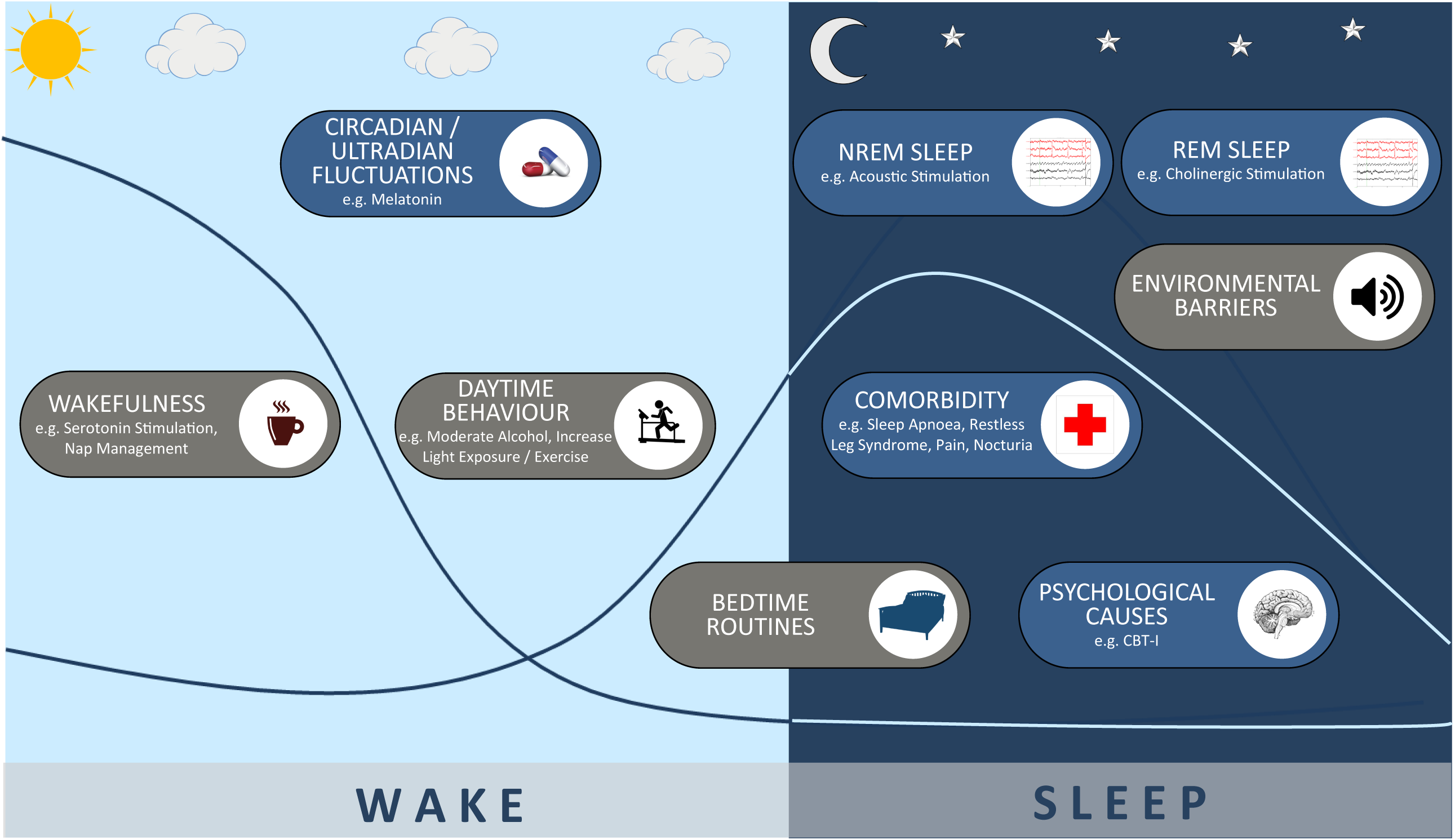
Theoretical interventional strategies to enhance sleep in people developing Alzheimer’s Disease

Here we systematically review the literature on sleep interventions for people meeting established criteria for either Mild Alzheimer’s Disease or Mild Cognitive Impairment. We focus specifically on AD rather than other subtypes of dementia as there are specific mechanisms proposed to link sleep disturbance to AD. In the absence of trials of sufficient length or size to establish the efficacy of sleep disturbance treatment on AD disease progression, we first review studies that measure the effect of intervention on sleep as an initial step in the translational pathway to slowing AD progression. This review does not explore the effect of interventions on specific sleep phenotypes due to the paucity of data. Improving sleep regardless of presence or absence of sleep disorder could plausibly impact on AD. Therefore inclusion and exclusion of any underlying sleep disorders in study design was noted, permitted and discussed.

In order to assess the degree to which different avenues for treating sleep in early AD have been explored, we, a priori, created a logic model to incorporate different possible intervention targets (**Figure 1**). We then compare the scope of existing studies with the model to guide where gaps might lie. The 9 targets included; promotion of wakefulness (to build sleep pressure), optimising circadian fluctuations, improving bed time routines (to remove barriers to sleep), removing overnight environmental barriers to sleep, enhancing Non-REM (NREM) sleep, improving REM sleep, treating psychological causes of insomnia, optimising day time behaviour to promote sleep at night and treating comorbidities that interrupt sleep.

## METHODS

The review followed recommendations by the Preferred Reporting Items for Systematic Review and Meta-Analyses Statement (PRISMA) (Moher et al., 2009). The protocol was registered in Prospero database (registration number CRD42019126320).

### Eligibility Criteria

All peer reviewed articles written in English were eligible for inclusion. The following criteria were used.

#### Participants

Inclusion criteria:

1. Adults aged greater than 18 (limit set to avoid excluding studies in genetic dementias); and
2. Male or Female; and
3. At least 75% of the studied group meet the following criteria:

a. Satisfies established diagnostic criteria for MCI e.g. (Albert Criteria, Peterson Criteria) or would be expected to meet these criteria if the study was conducted before 1999. Or:
b. Satisfies established diagnostic criteria for AD dementia e.g. (DSM-IV, ICD-10, ICD-11, NINCDS-ARDA, DSM-V) or would be expected to meet these criteria if study conducted before 1984.

Exclusion criteria:

1. In order to focus on mild AD and MCI we excluded studies where a majority of the studied group is likely to have Moderate to Severe AD dementia as evidenced by:

a. Mini Mental State Examination (MMSE) less than 20; Or:
b. Clinical Dementia Rating (CDR) greater than or equal to 2; Or:
c. Equivalent measure.

#### Exposure

Interventions of interest include any pharmacological or non-pharmacological treatment / technique primarily utilised to improve the duration or quality of sleep.

#### Comparator

The studied intervention must be compared with at least one other intervention, non-exposure or placebo.

#### Outcome

Sleep outcomes e.g. total sleep time, sleep efficiency or sleep latency measured by an appropriate validated sleep outcome measure.

#### Study Design

Eligible studies included comparison study designs e.g. systematic reviews and meta-analyses. Randomised Controlled Trials (RCTs), group trial designs e.g. case-control, cross-sectional or prospective cohort studies as well as multiple base studies were also eligible. Case reports, abstracts and grey literature were excluded.

### Search Strategy

Medical Subject Headings (MeSH) index terms were used together with free text to capture relevant results. Preliminary research was undertaken to identify relevant synonymous terminology and refine the search strategy. The full list of search terms is provided in **Appendix 1**. The following electronic databases were searched: The Cochrane Library, MEDLINE (1946 to present), EMBASE (1974 to present), CINAHL Plus (1937 to present), British Nursing Index (1994 to present) and PsycINFO (1806 to present) and the trial registry WHO ICTRP. Citations of the included papers were hand searched for additional studies. The search was undertaken on 19^th^ August 2019.

### Study Selection

Search results were merged using reference management software and duplicate records of the same report removed. A sample of 10% of the total titles were reviewed by each reviewing team member to ensure consensus in decision making. Remaining titles were reviewed individually by a member of the reviewing team in order to exclude studies obviously not meeting criteria.

Abstracts of the retained references were examined against inclusion and exclusion criteria with a further sample of 10% reviewed by each reviewing team member to ensure consensus. The full texts of remaining articles were checked against inclusion / exclusion criteria by two members of the review team. In the process of the search, the reference lists of any identified relevant review articles were hand searched to ensure capture of references not identified on the electronic databases.

Retained studies were recorded in a spreadsheet and included study characteristics, details of the patient/population group, intervention and primary outcomes. Where required, correspondence with investigators was attempted in order to gain further study data or to clarify study eligibility. Discrepancies were resolved through discussion with the first author (JB) in consensus with the reviewing team.

### Data Extraction

Data extracted from the retained articles (**Table 1**) included: Participant Characteristics (No. of participants, country, gender, age), Condition / Diagnosis, Sleep Co-Morbidity, Intervention(s) Studied, Intervention Dosage / Duration, Control Group, Validated Outcome Measure Used, Sleep Parameters Measured, Data on Efficacy, Data on Complications.

**Table 1.** Individual Study Characteristics

| Author/<br>Year | Condition /<br>Diagnosis | Sleep Co-Morbidity<br>Status | Country | No. Male<br>(Female) | Mean<br>Age<br>(SD) | Study Type | Interven-<br>tion | Treatment<br>Dosage / Duration | Control | Validated<br>Outcome<br>Measure | Sleep Parameters<br>Evaluated |
| --- | --- | --- | --- | --- | --- | --- | --- | --- | --- | --- | --- |
| Cassidy-Eagle et<br>al., 2018a<br>and<br>2018b* | MCI | Insomnia<br>(DSM IV<br>Criteria) | United<br>States | 28 (24) | 89.4 | Randomised<br>Controlled<br>Trial | Cognitive<br>Behavioural<br>Therapy -<br>Insomnia | Six 1-hour sessions | Active control<br>class | Wrist Actigraphy<br>ISI | TST, SE, WASO, SL<br>ISI Total |
| Cooke et<br>al., 2006 | Mild to<br>Moderate AD | Sleep / OSA<br>Co-Morbidities<br>Excluded | United<br>States | 76 (25) | IG1 77.8<br>(7.4)<br>IG2 77.6<br>(11.5)<br>IG3 79.8<br>(8.3)<br>CG 78.3<br>(6.2) | Prospective<br>Study | Donepezil<br>Galantamine<br>Rivastigmine | On stable dose of<br>AChE Inhibitor for 2<br>months | Non<br>Medication<br>Arm | Polysomnography | TST, SE, WASO, SP,<br>%REM, %N1, %N2,<br>%N3/N4, TIB, PLMI, AI,<br>AH |
| Cooke et<br>al., 2009 | Mild to<br>Moderate AD<br>with OSA | OSA only | United<br>States | 52 (13) | 77.8 (7.3) | Randomised<br>Controlled<br>Trial | CPAP | Therapeutic CPAP for<br>3 weeks in<br>intervention arm<br>Therapeutic CPAP for<br>3 weeks in both arms | Placebo CPAP | Polysomnography | TST, SE, WASO, SL, SP,<br>%REM, %N1, %N2,<br>%N3/N4, TIB, AI |
| Cooke et<br>al., 2009 | Mild AD with<br>OSA | OSA only | United<br>States | 10 (3) | 75.7 (5.9) | Prospective<br>Study | CPAP | Continued use' over<br>approximately 3 years | Non<br>Intervention<br>Arm | PSQI<br>ESS<br>FOSQ | PSQI Total<br>ESS Total<br>FOSQ Total |
| Jean-Louis<br>et al.,<br>1998 | MCI and Mild<br>AD | Self reported<br>sleep-wake<br>disturbance | United<br>States | 10 (6) | 68.8 | Double-Blind<br>Crossover<br>Trial | Melatonin | 6mg nocte for 10 days | Placebo Arm | Wrist Actigraphy | TST, SE, WASO, SL,<br>Transition, TWT |
| Markowitc et al., 2003 | Mild to Moderate AD | Mixed Population +/- Sleep Disorder | Canada<br>New Zealand<br>South Africa<br>UK<br>USA<br>Belgium | 261 (157) | CG 74.6 (7.6)<br>IG 75.3 (7.5) | Randomised Controlled Trial | Galantamine | 12mg bd | Placebo Arm | PSQI | PSQI TST, SE, SL, Total, Daytime Dysfunction, Sleep Disturbances, Subjective Sleep Quality |
| Moraes et al., 2006 | Mild to Moderate AD | Moderate / Severe Sleep Disorders excluded | Brazil | 35 (24) | IG 77.4 (6.6)<br>CG 74.5 (9.8) | Randomised Controlled Trial | Donepezil | 5mg od for 1 month increasing to 10mg od | Placebo Arm | Polysomnography<br>EEG Spectral Analysis | TST, SE, SL, %REM, %N1, %N2, %N3/N4, PLMI, AH, REM Characteristics |
| Naharci et al., 2015 | AD and Mixed Dementia | Mixed Population +/- Sleep Disorder | Turkey | 78 (47) | IG1 81.5 (6.9)<br>IG2 80.9 (7.4)<br>IG3 79.1 (7.7)<br>CG 77.4 (66.3) | Prospective Study | Donepezil<br>Galantamine<br>Rivastigmine | 5-10cm<sup>2</sup> od<br>5-10mg od<br>8,16,24mg od | Healthy Population | PSQI | PSQI Total |
| Naismith et al., 2018 | MCI | Primary Sleep Disorder excluded | Australia | 37 (19) | IG1 69.4 (9.5)<br>CG 70.0 (8.8) | Randomised Controlled Trial | "Sleep Well, Think Well" Group Program | 8 week group program (Four 1hr face to face sessions and four telephone session) | Non-directive Control Group | PSQI<br>ESS<br>Wrist Actigraphy | TST, SE, WASO<br>PSQI Total<br>ESS Total |
| Papalambros et al., 2019 | MCI | Circadian Rhythm Disorders / OSA excluded | United States | 9 (5) | 72 | Randomised Crossover Trial | Phase Locked Loop Acoustic Stimulation | Parameters for stimulation as per previous established algorithm (Santostasi et al., 2016) | Sham Stimulation | Polysomnography | TST, SE, WASO, SL, %REM, %N1, %N2, %N3/N4, AI, Spindle Characteristics |
| Petit et al., 2003 | Mild to Moderate AD | Mixed Population +/- Sleep Disorder | Canada | 8 (3) | 61.9 | Double-Blind Crossover Trial | Tetrahydroaminoacridine (THA) | 50mg od for 1 week<br>75-100mg od for subsequent week | Placebo Period | Polysomnography | SE, SL, %REM, %N1, %N2, %N3/N4, REM Characteristics |

\*2 separate papers written by Cassidy-Eagle identified reporting on same study data. Analysis was focussed on paper reporting in more detail on sleep outcome measures to avoid over-representation of data. Papers reported separately in quality appraisal and risk of bias assessment for completeness.
Key: AH – Apnoea Hypopnoea Index, AI – Arousal Index, CG – Control Group, DSM – Diagnostic and Statistical Manual, ESS – Epworth Sleepiness Scale, FOSQ – Functional Outcomes of Sleep Questionnaire, IG – Intervention Group, ISI – Insomnia Severity Index, NINCDS-ADRDA - National Institute of Neurological and Communicative Disorders and Stroke and the Alzheimer's Disease and Related Disorders Association, OSA – Obstructive Sleep Apnoea, PLMI – Periodic Limb Movement Index, PSQI – Pittsburgh Sleep Quality Index, REM – Rapid Eye Movement, SE – Sleep Efficiency, SL – Sleep Latency, TIB – Time in Bed, TST – Total Sleep Time, TWT – Total Wake Time, WASO – Wake After Sleep Onset

### Quality Assessment

Methodological quality of eligible studies was evaluated using the Joanna Briggs Institute Checklist for Quasi-Experimental and Randomized Controlled Trials (Tufanaru et al., 2017). Independent evaluation of quality was undertaken by two reviewers (SH and JB) and consensus reached jointly.

Risk of bias for all included studies was also assessed using the Cochrane Collaboration’s Tool for Assessing Risk of Bias (Higgins et al., 2019). Two reviewers (SH and JB) undertook this analysis independently before reaching joint consensus.

### Data Synthesis and Analysis

Due to substantial heterogeneity in outcome measures no meta-analysis of quantitative data was appropriate. Therefore a narrative synthesis of included studies was performed focusing on population characteristics, interventions utilised and outcomes. The main focus of analysis was on the performance of the intervention assessed against a comparison intervention or control.

## RESULTS

### Included Studies

The search strategy identified 25,148 unique records alongside an additional 36 identified through bibliographic search. 660 abstracts and subsequently 284 full text articles were screened for eligibility with a total of 12 articles selected for inclusion, (see PRISMA flowchart - **Figure 2**). There were two predominant reasons for exclusion, 1) ineligible study type e.g. intervention not analysed against an alternative or control, 2) ineligible population e.g. due to severity of dementia. The 12 articles identified reported on data from 11 separate studies. Attributes of individual included studies are listed in **Table 1**.

**Figure 2.**
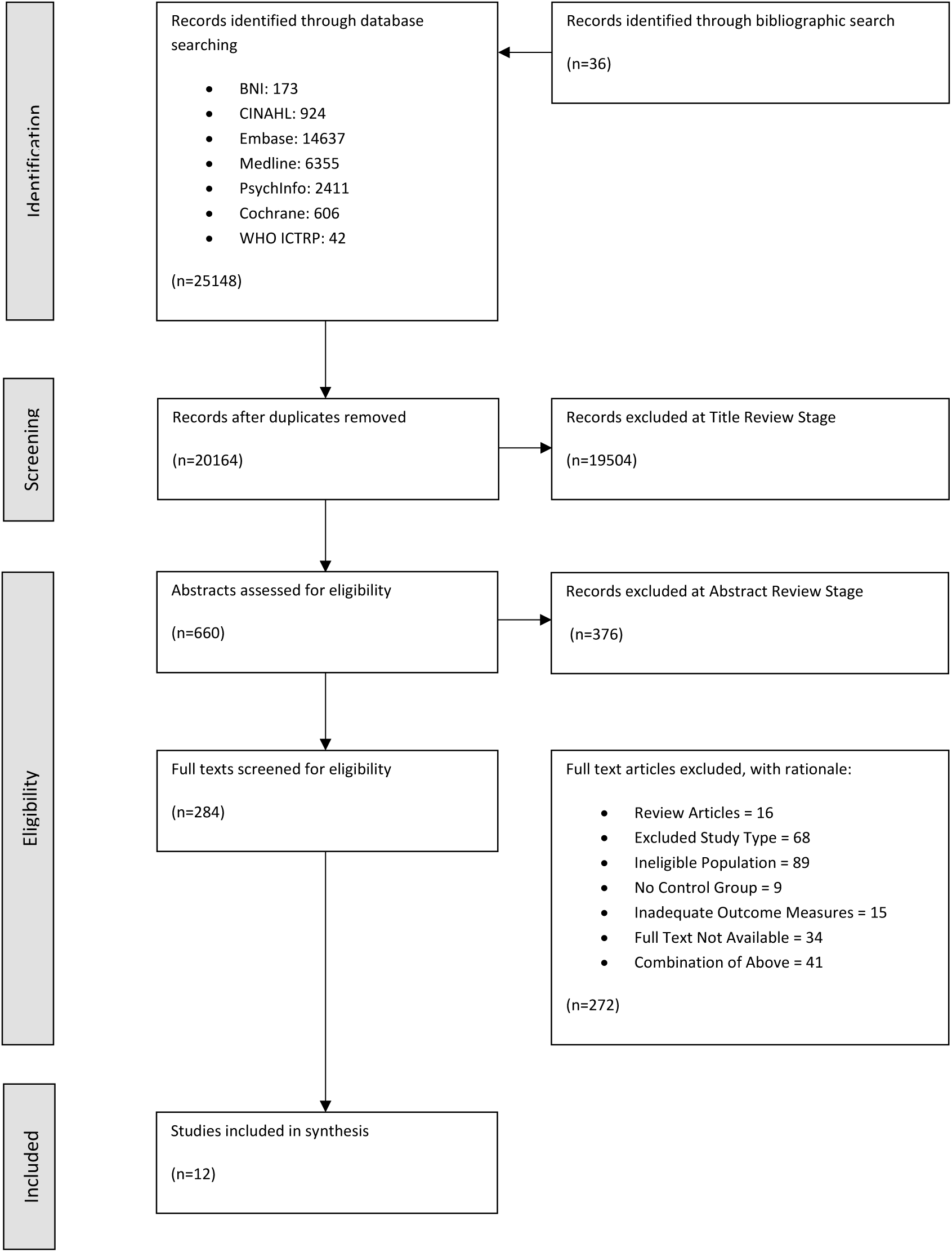
PRISMA Flowchart

### Participants and Comparison Measures

A summary of combined included study characteristics is shown in **Table 2**.

**Table 2.** Combined Included Study Characteristics

|  |  |
| --- | --- |
| **Demographics** |  |
| Total Participants | 602 |
| Mean Age | 76.3 |
| % Female (Male) | 54.1 (45.9) |
| **Diagnosis and Severity** |  |
| Cognitive Diagnostic Criteria Utilised | NINCDS-ADRDA (n=7)<br>Peterson MCI Criteria (n=2)<br>Albert Criteria (n=1)<br>DSM-IV (n=1) |
| Assessment of Dementia Severity Tool | MMSE (n=8)<br>MOCA (n=1)<br>CDR (n=1)<br>Not reported (n=1)* |
| Baseline Cognitive Scores by Tool | Mean MMSE 20.56/30 (n=7)<br>Median MMSE 28/30 (n=1)<br>Mean MOCA 23.64/30 (n=1)<br>Mean CDR 1.35 (n=1) |
| Sleep Co-Morbidity Status | Sleep Disorders Included :-<br>Mixed Population +/- Sleep Disorder (n=3)<br>OSA (n=2)<br>Insomnia by DSM V Criteria (n=1)<br>Sleep Disorders Excluded :-<br>All (n=2)<br>Self-Reported Sleep-Wake Disturbance (n=1)<br>Moderate-Severe Sleep Disturbance (n=1)<br>Circadian Rhythm Disorders and OSA (n=1) |
| **Comparison** |  |
| Methodology Used | Placebo Groups (n=3)<br>Active Non-Pharmacological Control Groups (n=2)<br>Non-Treatment Arms (n=2)<br>Placebo Crossover Designs (n=3)<br>Healthy Population Group (n=1) |
| **Outcome** |  |
| Measurement Tools | Polysomnography (n=5)<br>Wrist Actigraphy (n=4)<br>PSQI (n=4)<br>ESS (n=2)<br>FOSQ (n=1)<br>ISI (n=1) |
| Outcome Measure (Tool Utilised) | TST (Polysomnography n=4, Actigraphy n=3, PSQI n=1)<br>SE (Polysomnography n=5, Actigraphy n=3, PSQI n=1)<br>WASO (Polysomnography n=3, Actigraphy n=3)<br>SL (Polysomnography n=4, Actigraphy n=2, PSQI n=1)<br>% REM, N1, N2, N3/N4 Sleep Duration (Polysomnography n=5)<br>AI (Polysomnography n=3)<br>PSQI Total (PSQI n=4)<br>Other e.g. PLMI, AH, Transition, TWT, Spindle Characteristics and REM Characteristics (Polysomnography n≤2) |

\* Diagnosis of MCI made utilising Peterson Criteria

### Outcome Measurement and Outcomes

As per **Table 2**, studies utilised a wide range of measurement tools with broad and heterogeneous primary and secondary outcome measures.

Directly comparable group data across key outcome measures was therefore sparse. TST in minutes and SE was provided in only 75 participants in intervention groups and 71 participants in control groups across three studies and for nine patients in a crossover design. Surrogate measures of TST and SE were provided by one study utilising PSQI in 127 intervention participants and 121 control participants and 124 intervention participants and 115 control participants respectively. Structural sleep % by stage N1,N2,N3/N4 and REM group means were provided in 108 patients in intervention groups and 55 in control groups across three studies and in 21 participants across 2 crossover studies. Actual Total PSQI Scores were provided in 198 intervention participants and 152 control groups across three studies.

### Adverse Events

No significant adverse events were reported across included studies. Donepezil was reported by Moraes et al (2006) to cause mild and transitory side effects including headache and nausea in 3 patients in an intervention of group of 17.

### Quality Appraisal and Risk of Bias

Quality appraisal was undertaken independently by two reviewers (JB and SH). Inter-rater reliability calculated using Cohen’s Kappa Coefficient was 0.823 [95% CI 0.727-0.919] indicating strength of agreement to be ‘very good’. For full tabulated results see **Figures 3–5**. The overall quality of included studies as measured by the appropriate Joanna Briggs Quality Appraisal Tool was variable. Multiple limitations were identified in one study, however this was partly due to inherent difficulties in concealment and blinding associated with utilising a psychotherapeutic intervention. One further study had 3 limitations identified (Cooke et. al 2006), two studies had two limitations identified (Petit et. al 1993 and Naismith et al 2018). The remaining seven studies had 0 or 1 limitation.

**Figure 3.**
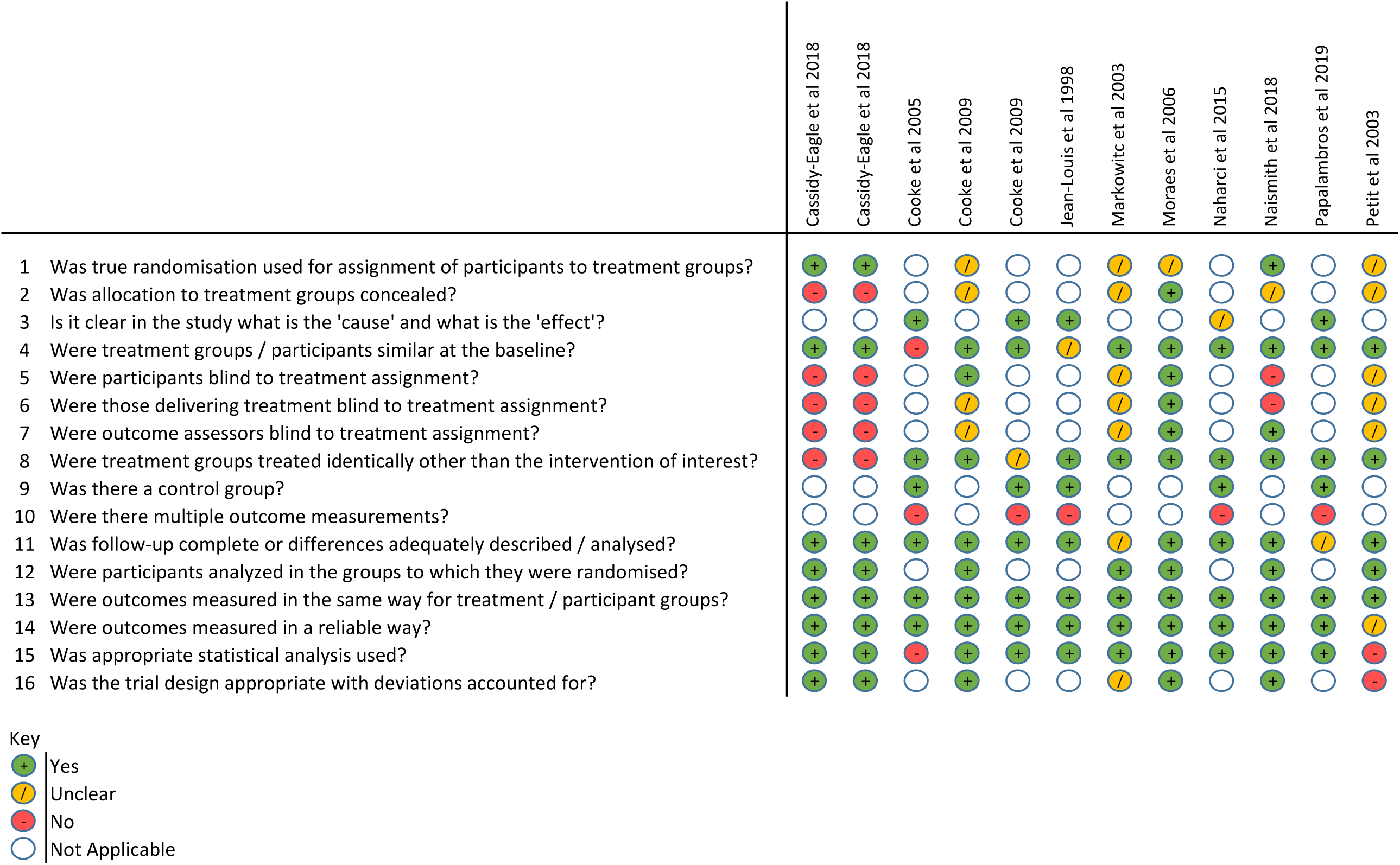
Quality Appraisal using Joanna Briggs Institute Tools by Study

**Figure 4.**
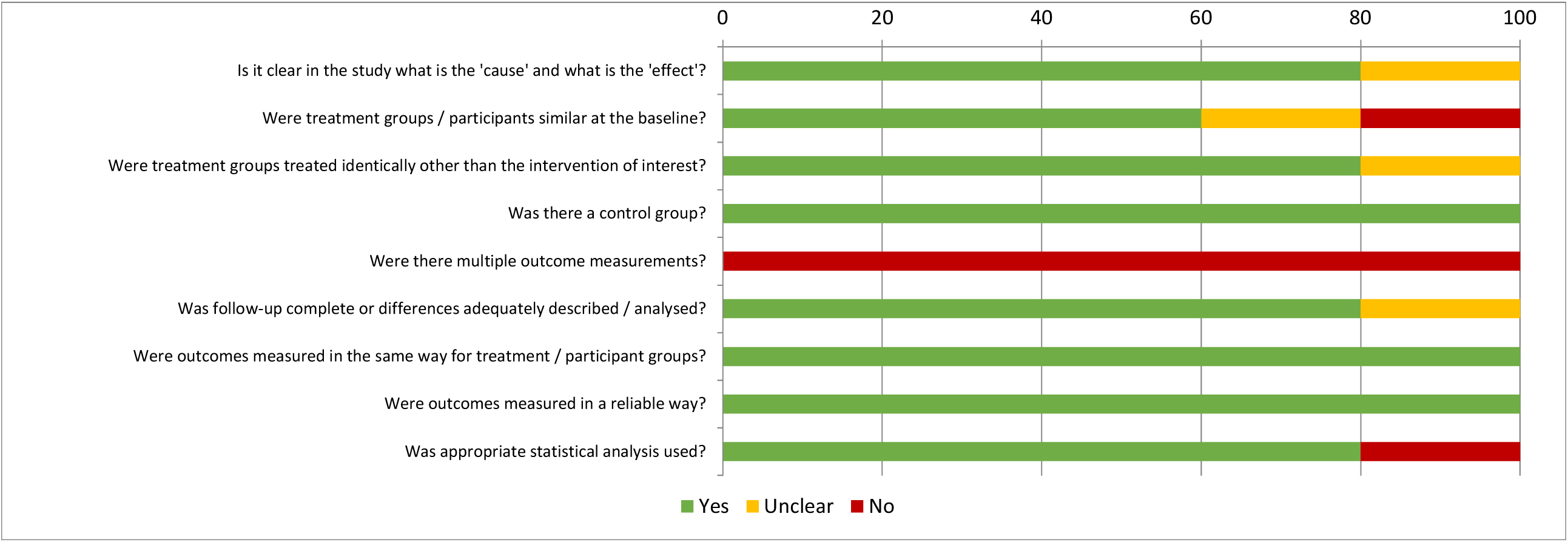
Overall Quality Appraisal by Joanna Briggs Institute - Quasi Experimental Tool

**Figure 5.**
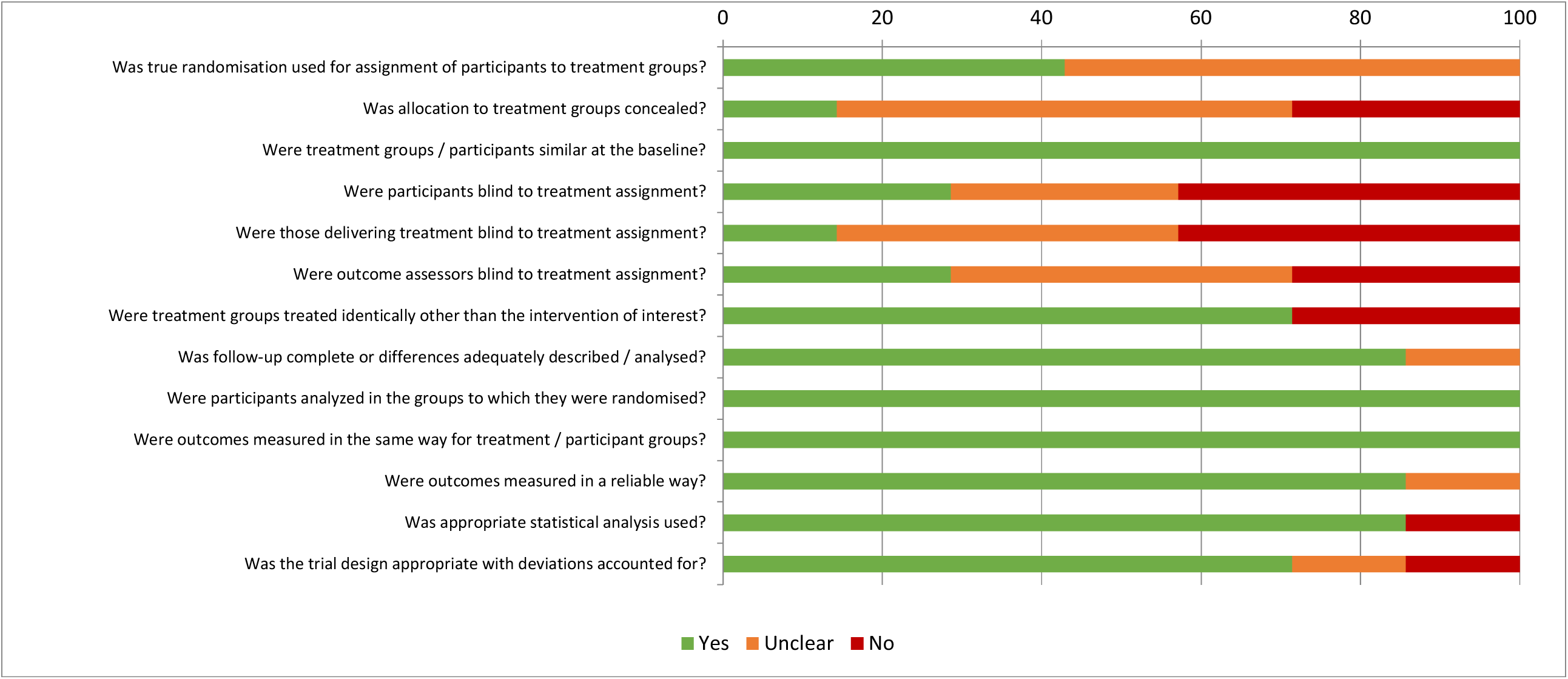
Overall Quality Appraisal by Joanna Briggs Institute – Randomised Controlled Trial Tool

Risk of Bias analysis using the Cochrane Collaboration Tool was also undertaken by two reviewers (JB and SH). Inter-rater reliability calculated with Cohen’s Kappa Coefficient was 0.751 [95% CI 0.631-0.870] indicating ‘substantial’ strength of agreement. High risk of bias in at least one area was identified in nine included studies with six identified as being at ‘high risk’ in multiple domains. For full results see **Figure 6**.

**Figure 6.**
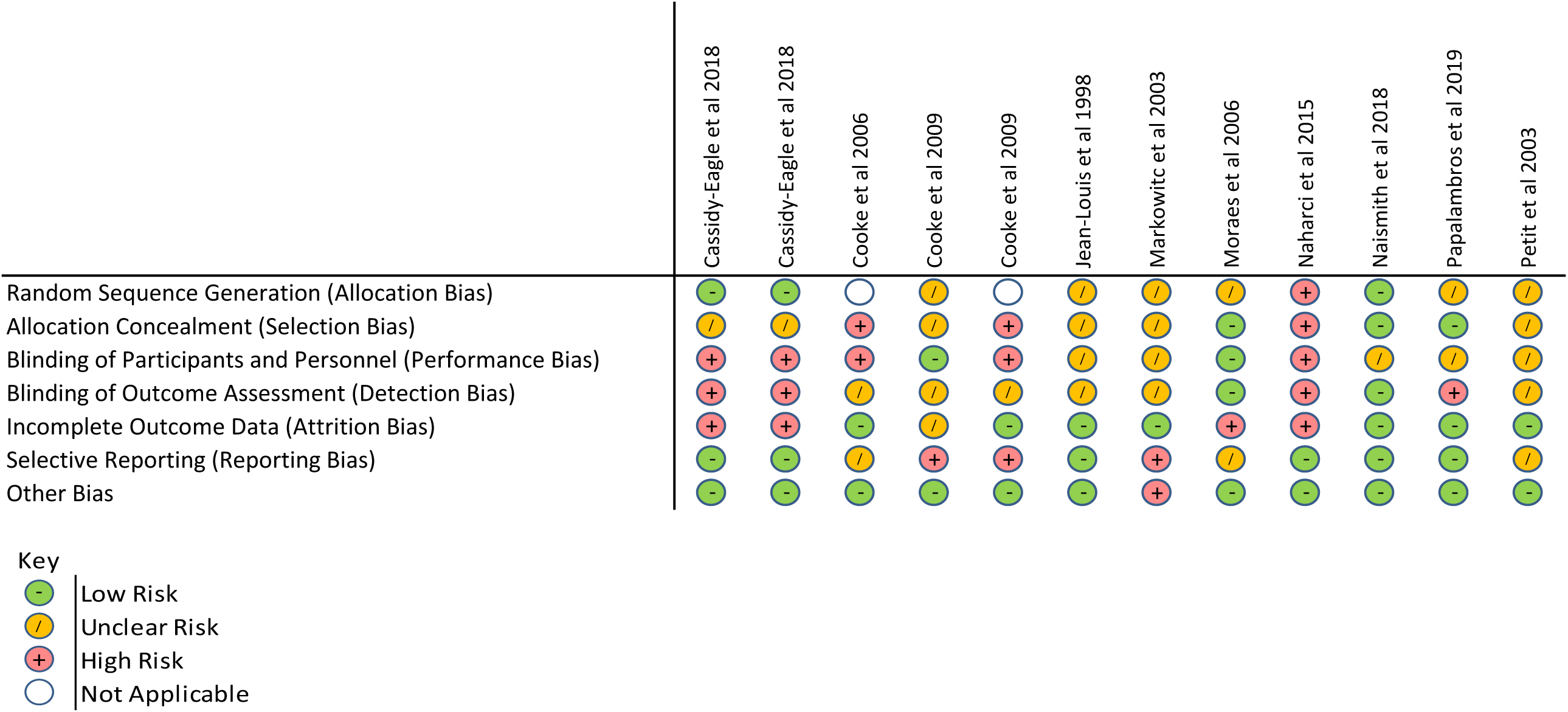
Cochrane Collaboration Tool for Assessing Risk of Bias

### Interventions

Overall we found trials in five of the nine intervention domains we have identified in **Figure 1**. None were large randomised controlled trials powered to find clinical benefit. Most were exploratory, pilot or feasibility trials. There was an over-representation of trials that used cholinesterase inhibitors to enhance sleep and there were no trials that aimed to modify day-time behaviour, promote wakefulness, optimise bedtime routines or remove environmental barriers to sleep. Results of individual trials within each potential intervention domain are as follows:

### Enhancing REM

#### Donepezil

Cooke et al (2006) compared donepezil against galantamine, rivastigmine and a group taking no Acetylcholine Esterase Inhibitor (AChEI). Participants had no prior sleep disorder and analysis was undertaken to test the hypothesis that AchEIs may increase REM sleep and nightmares. Polysomnographic analysis showed a significant decrease in Stage I sleep (p=0.01) and for those taking donepezil compared to those taking galantamine but no changes compared to other groups. There was also a significant increase in Stage II sleep (p=0.04) compared to the group taking no AChEI.

However this finding was not replicated when Moraes et al (2006) tested a similar hypothesis that Donepezil may increase REM sleep percentage and alter the REM sleep EEG power spectrum. Donepezil was directly compared against placebo in a randomized controlled trial of 35 participants with mild to moderate AD and without moderate or severe sleep disorders. No significant differences were found polysomnographically between donepezil and placebo for TST, SE, SL, REM SL or NREM sleep stage percentages. Donepezil was, however, found to significantly increase REM sleep percentage (p<0.01). Fast Fourier transformation of electroencephalogram (EEG) channel data as part of REM EEG Spectral Analysis also revealed significant decreases in overall theta band power and frontal delta and theta band powers after 6 months of donepezil compared with placebo (0.01≤p≤0.04).

Naharci et al (2015) compared total PSQI prospectively in 87 patients prescribed Donepezil, Galantamine or Rivastigmine and a control group not taking an AChEI with the rationale to evaluate the broad influences of AchEIs on sleep pattern. Study participants included those with and without underlying sleep disturbance. No significant change in total PSQI score was found across groups.

### Galantamine

Galantamine was found to have little or no impact on sleep architecture by Cooke et al (2006) in comparison with a group taking donepezil, rivastigmine and a group taking no AChEI. Markowitz et al (2003) undertook a post-hoc analysis of a RCT comparing a flexible dose regime of galantamine with placebo. It aimed to explore whether adverse sleep outcomes were associated with galantamine both in participants with and without underlying sleep disorders. Analysis of PSQI and Neuropsychiatry Inventory Sleep Disorder Items revealed no significant difference between groups or from baseline to post intervention. However more recently Naharci et al (2015) found in a non-randomised trial that galantamine when compared against donepezil, rivastigmine and a healthy control group significantly decreased total PSQI score (p=0.028). The highest improvement rate was in those with reported poor sleep quality.

### Rivastigmine

Rivastigmine was not associated with any change in sleep architecture according to Cooke et al (2006) although the authors acknowledge that their small sample size (n=8) is likely to have reduced the power to detect a significant effect. Naharci et al (2015) also found that rivastigmine decreased total PSQI score although this did not reach statistical significance (p=0.193).

### Tetrahydroamineacridine – (Tacrine -THA)

The centrally acting anticholinesterase inhibitor THA was compared against placebo by Petit et al (1993) in a small crossover trial (n=8) involving participants with mild to moderate AD. THA was hypothesised to alleviate sleep disturbance in this group and restore EEG desynchronisation during wakefulness and REM sleep. There were no differences in sleep architecture including SE, SL, SE and percentages of REM and NREM sleep on polysomnographic analysis in the THA vs placebo group. Spectral EEG Analysis by fast Fourier transformations utilising the ratio of power in slow (delta and theta) frequencies to fast (alpha and beta) frequencies revealed REM sleep EEG slowing in the frontal and parietoocciptal regions in the THA group compared to baseline (p<0.01).

To *summarise*, included studies showed that donepezil may have an impact on sleep architecture through altering the relative proportion of sleep stages, however the precise nature of this effect was not consistently demonstrated. Certainly it has not been shown to improve basic sleep parameters e.g. TST, SE and SL. Galantamine appears to have little effect on sleep architecture^31^ but may improve PSQI scores in a small non-randomised study^37^. Again we have found little in the way of replicated findings. Rivastigmine was not demonstrated to have any effect on sleep^31,37^. THA may reduce EEG REM sleep cortical slowing as measured by amplitude spectral EEG analysis^41^, although the clinical relevance of this is questionable in a largely withdrawn medication with no demonstrable positive effects on sleep.

### Optimising circadian fluctuations

#### Melatonin

A placebo controlled crossover trial of melatonin was conducted by Jean-Louis et al (1998) in 10 participants with MCI and Mild AD with self-reported sleep-wake disturbance either through difficulty in sleep initiation or frequent nocturnal awakenings. The study aimed to provide objective evidence of its effect on sleep, mood and cognition in the context of previously conflicting evidence.

Actigraphy revealed melatonin to have a statistically significant effect on sleep latency (melatonin group=14.7 mins [SD 7.21], placebo group=26.08 mins [SD 10.97], p=0.018). There was also a significant reduction in number of transitions from sleep to wakefulness (melatonin group=17.75 [SD 7.79], placebo group=31.20 [SD 11.37], p=0.011). There was no significant effect on SE, TST, WASO or phase shift in rest-activity pattern.

### Enhancing NREM

#### Phase Locked Loop Acoustic Stimulation

Nine participants with Amnestic MCI were recruited for a randomised crossover sham-controlled study conducted by Papalambros et al (2019) in order to examine the feasibility of acoustic stimulation during overnight sleep in enhancing Slow Wave Activity (SWA). Participants had no evidence of a circadian rhythm disorder or moderate-severe sleep apnoea. Acoustic Stimulation was delivered as per a previously established protocol^40^ for one night on two separate occasions seven days apart.

There were no statistically significant effects on basic sleep parameters e.g. TST, SE, SL, WASO or in sleep staging characteristics. Spectral analysis revealed that Slow Oscillation (SO) Activity and Slow Wave Activity (SWA) both increased during Stim ON intervals compared to Sham ON intervals by 15.1% (2.8) and 11.4% (2.9) respectively p<0.01. A similar decrease in SO and SWA was observed comparing Stim OFF with Sham OFF of 8.1% (3.3), p=0.040 and 6.6% (2.9), p=0.046 respectively. Within the same night, Stim ON appeared to increase SO by 22.2% (5.1), p=0.003 and SWA by 17.9% (4.12), p=0.004 when compared with Stim OFF. However overall, there was no difference in mean NREM SWA between stim and sham conditions. There were no statistically significant effects on spindle characteristics or on N2,N3 duration.

### Treating Co-Morbidities

#### Continuous Positive Airway Pressure (CPAP)

Patients with Obstructive Sleep Apnoea (OSA) but no prior diagnosis of a sleeping disorder and AD were recruited to a RCT by Cooke et al (2009). The effect of CPAP on sleep was compared with placebo CPAP using polysomnography. Those receiving therapeutic CPAP after one single night had significantly decreased Stage I sleep (p=0.04) and significantly increased Stage 2 Sleep (p=0.02). There were no changes in % REM, Stage III, IV Sleep, WASO, SP, TIB, SE or SO in either group. Paired analysis after 3 weeks showed that there were decreases in mean WASO (p=0.005), TIB (p=0.002), SP (p<0.001), Arousals per Hour (p=0.005) and % Stage I Sleep (p=0.001) and a significant increase in Stage III Sleep (p=0.006). In summary CPAP was reported to deepen the level of sleep immediately and later lead to more consolidated sleep with fewer night-time awakenings and improved SE.

Cooke et al (2009) also undertook analysis of the potential longer term effects of CPAP on sleep comparing 5 patients who had continued CPAP (CPAP+) following a previous randomized trial against 5 patients who had discontinued CPAP (CPAP-) approximately 3 years after the initial study. The CPAP+ group were found to have a significant statistical improvement in PSQI while the CPAP-group showed a statistical deterioration (effect size = 1.8, p<0.05). At baseline neither CPAP+ nor CPAP-groups had evidence of daytime somnolence as measured by the Epworth Sleepiness Scale (ESS). However at follow-up, ESS in the CPAP-group had deteriorated in keeping with significant daytime somnolence (effect size = 0.8) whilst ESS in the CPAP+ group remained stable compared to baseline.

### Treating psychological factors causing insomnia

#### Cognitive Behavioural Therapy – Insomnia (CBT-I)

Cassidy-Eagle et al (2018) employed a two-arm RCT to explore effects of CBT-I on a sample of 28 participants with MCI and a DSM-IV diagnosis of insomnia. A standard CBT-I protocol over 6 weeks was adapted for participants with MCI through a reduction in content covered, inclusion of brief, focused rationales for treatment, time allowed for review and repetition of content as well as reminder / troubleshooting calls between intervention sessions. It was compared against an ‘Active Control Nutrition Class’ with identical in-person and phone contact hours which provided learning modules delivered by a dietician covering the impact of diet on aging. These adaptations to standard CBT were hypothesised to improve both objective actigraphy variables and self-reported insomnia symptoms through the ISI.

CBT-I was shown to have a highly significant desirable effect on 4 out of 5 outcome variables. ISI decreased from 15.29 (SD 2.33) to 3.25 (SD 2.05) in the intervention group, SL from 11.03 (SD 11.69) to 1.93 (SD 3.57), WASO from 104.24 (SD 38.21) to 46.95 (SD 25.09) and SE increased from 0.792 (SD 0.0744) to 0.8804 (SD 0.0719). Multilevel regression analysis revealed these outcomes as statistically significant with large effect sizes ISI (p<0.001 Cohen’s d^2^ −4.22), SL (p<0.001 Cohen’s d^2^ − 1.73), WASO (p<0.001 Cohen’s d^2^ −2.32), SE (p<0.001 Cohen’s d^2^ 1.89). TST, however, appeared to decrease from 436.61 (SD 51.53) to 379.50 (SD 75.13) in the intervention group, also finding statistical significance with a large effect size (p=0.02 Cohen’s d^2^ −2.32)

#### “Sleep Well – Think Well” Group Program

Naismith et al (2018) evaluated the “Sleep Well – Think Well” Program in a pilot RCT of 37 participants with MCI but without underlying sleep disorders. The program involved provision of five booklets, specifically a relaxation booklet, anxiety and worry booklet, healthy sleep practices and intervention booklet, mindfulness booklet and a booklet detailing helpful / unhelpful thoughts associated with sleep initiation and maintenance to both active and control groups. The active intervention group received four face-to-face sessions consisting of psychoeducation, introduction of a guided individualised treatment plan, monitoring of the treatment plan and a summary and consolidation session. Fortnightly compliance monitoring telephone sessions were also conducted. The control group received fortnightly calls to discuss handbook content but no directive, supportive input as above.

There was evidence of improved subjective sleep through a statistically significant reduction of mean total PSQI score in the intervention group (Mean change −2.4 p=0.023 Cohen’s d=0.83). Analysis of individual PSQI components showed statistically significant improvement in SE (p=0.049) and daytime dysfunction (p=0.015). There were no statistically significant changes in objective actigraphy parameters including TST, SE and WASO.

## DISCUSSION

### Overall Findings

Comprehensive literature search revealed a relative paucity of robust data evaluating sleep interventions and their efficacy in a population with mild AD or MCI. The majority of studies were of small scale and many subject to potential QA limitations and bias. In all of the potential theoretical domains for intervention (**Figure 1**), there are potential strategies that have not yet been tried. Additionally with the possible exception of the ‘Sleep Well, Think Well’ trial most studies focussed on one intervention as opposed to a multi-modal approach.

The most widely studied intervention were the AChEIs likely due to a combination of theoretical, clinical and pragmatic reasons. Early neurochemical theories of sleep hypothesised Acetylcholine playing a primary role in wakefulness and REM sleep (Jouvet, 1972), further strengthened by later work underlying the muscarinic M2 receptor’s role in potentiating REM sleep (Baghdoyan & Lydic, 1999; Watson et al., 2012). Clinically, donepezil has been associated with vivid dreams and insomnia with higher incidences vs placebo reported in multiple studies (Burns et al., 1999; Pratt et al., 2002; Rogers et al., 1998) which alongside their common use in those with early cognitive impairment may have driven further research.

The results of trials included here did not reveal a consistent benefit of AChEIs on sleep and it seems unlikely this class of medications will be pursued further to benefit sleep. The one exception to this could involve new technology for drug delivery to modulate available acetylcholine to better mimic physiological fluctuations in cholinergic activity. Given we know that cholinergic synapses are involved in learning while awake (Huerta & Lisman, 1993) and less so during sleep, perhaps more refined temporal target (e.g. daytime only) could improve nocturnal sleep and memory consolidation. We generally noted that there were no identified studies where a daytime-only pharmacological intervention, promoting wakefulness or attention, was utilised as a tool to enhance night-time sleep, and this could be a useful future approach.

In a population with OSA and mild AD /MCI, CPAP does appear to be an effective intervention with significant positive effects demonstrated both in sleep architecture and key sleep parameters. Sleep apnoea affects a large proportion of people aged over 50 (Heinzer et al., 2015), therefore treating sleep apnoea holds great promise to improve quality of life and delay AD progression. Sleep apnoea is sufficiently common that screening should be a routine part of clinical practice in memory clinics - we already know that patients can benefit through treatment of sleep apnoea regardless of whether they have AD. The possibility of treating AD is an added benefit. We also propose that screening and, if required, treatment of sleep apnoea should occur before entry into AD clinical trial of any type. Unless we exclude the impact of sleep apnoea on AD progression, we may be clouding benefit from other therapeutic approaches.

Two studies explored the effect of tailored psychotherapeutic programs designed on principles of Cognitive Behavioural Therapy. Outcomes were mixed, with Naismith et al (2018) reporting a substantial improvement in subjective sleep quality but observing no significant change in objective actigraphy measures. In contrast, Cassidy-Eagle et al (2018) reported significant improvement in 4 out of 5 objective outcome measures including SL, WASO and SE as well as significant improvements in subjective quality (ISI). Of note, however, there were potentially significant differences between the two populations with those recruited to the Cassidy-Eagle (2018) study requiring a DSM-IV diagnosis of Insomnia in contrast to the Naismith et al (2018) study. Critically, both studies highlight the feasibility of this approach in MCI.

Melatonin appeared to statistically reduce sleep latency and the number of overnight transitions from sleep to wakefulness but in an extremely small study of 10 participants. Other approaches to circadian rhythm modification have not been systematically explored despite evidence that circadian rhythms are disturbed in AD – although we note protocols suggesting trials are underway (Falck et al., 2018).

Phase locked acoustic stimulation appeared to have no effect on basic sleep parameters or sleep-stage architecture but did appear to increase SWA and SO whilst ‘ON’ without increasing its mean overall when compared to a sham comparison group.

The lack of available evidence was surprising given the broad range of pharmacological and non-pharmacological sleep interventions available for use in the general population. Striking omissions include the most common group of pharmacological interventions for insomnia (Z-hypnotics such as zopiclone) which appear to have no evidence base in this group.

There were multiple interventions such as vitamin B12, risperidone, acupressure, bright light therapy, transcutaneous electric nerve stimulation (TENS) and music interventions that were not included in our review either due to the use of unvalidated sleep outcome measures or because there was no comparison group to allow for meaningful conclusions to be drawn. In three cases the diagnoses of mild AD /MCI could not be confirmed to have been made using recognised criteria.

Lifestyle modification was explored only in the context of CBT-I and multi-modal approaches rather than in isolation. In established mild-moderate AD dementia there are concerns about cognitive limitations that might prevent behavioural change. However at the earlier stages, cognitive function often still allows significant adaptation of lifestyle and motivation can be high. Furthermore, pharmacological interventions carry with them potential toxicity and the burden of polypharmacy. For these reasons, behavioural interventions may well be a fruitful route for further exploration.

The need for effective sleep interventions is highlighted through the recent emergence of plausible mechanisms through which poor sleep could result in AD progression. Beta amyloid is one of the two pathognomonic changes of AD. Levels of soluble Amyloid Beta fluctuate diurnally in both mice and humans showing the crucial role that sleep plays in its clearance (Huang et al., 2012). Amyloid Beta 42 levels have been shown to be significantly increased after a single night of sleep deprivation in healthy adults (Ooms et al., 2014) and slow oscillations are temporally linked to CSF flow (Fultz et al., 2019). Chronic sleep deprivation accelerates accumulation of soluble beta amyloid into insoluble amyloid plaques in two mouse models (Kang et al., 2009). A cascade is envisaged in which poor sleep disrupts the clearance of soluble amyloid, leading to plaque deposition. This precipitates further plaque formation through recognised positive feedback loops (Mandrekar-Colucci et al., 2012). Slow wave sleep (Non-REM) declines as part of normal ageing and its reduction is likely to be a pervasive risk factor for AD progression. It should be noted that we do not yet know whether improving sleep will slow or prevent accumulation of amyloid. Nevertheless specific targeting of slow wave sleep through medications, sound technology or other approaches is one particularly interesting avenue for intervention to improve cognition and possibly slow progression of AD pathology.

### Study Strengths and Limitations

This is the first review we know of to focus on sleep intervention in MCI and Mild AD. A comprehensive electronic database search protocol was performed with a wide range of potential sleep interventions sought. Additionally, we summarized findings on multiple dimensions of sleep outcome data. A process for rigorous justification of all subsequent decisions in terms of inclusion/exclusion was utilised. As with the inclusion criteria for AD dementia/MCI, attempts were made to contact authors in cases of ambiguity.

A significant challenge in this review was in formulating inclusion criteria to ensure that studies involving important, clinically relevant interventions were included but also ensuring that studies with an overly heterogeneous population whose findings may not be applicable to our population of interest were excluded. A threshold of 75% was chosen for participants to have an established diagnosis of AD dementia or MCI. It was further decided to exclude any study where the majority of participants have moderate or severe dementia. Naturally these cut-offs could be criticised either for diluting the population of interest or for failing to allow for inclusion of relevant studies. We therefore cannot exclude the possibility that there may be important findings left unreported. Included studies reported on a range of diverse outcomes, which whilst individually valid, made comparison across studies challenging. Direct comparison was further hindered by variations in study designs permitting or excluding the presence of underlying recognised sleep disorders amongst participants. Ideally, the effect of interventions on specific sleep phenotypes would have been reviewed, however unfortunately this information was not reported on the scale to allow for meaningful comparison. Nonetheless our decision to include studies regardless of phenotype could be argued to increase real-world applicability to the clinical population and is not contrary to the central hypothesis that an improvement in sleep regardless of baseline may positively impact on AD progression. Quality appraisal scores across included studies were variable and those included were largely subject to at least one domain of significant bias thereby potentially compromising their findings. This though, remains consistent with our assertion that there is a paucity of reliable, large-scale data for this population. For this present review, the risk of publication and language bias for current literature is also recognised.

### Clinical Implications and Future Direction

The lack of rigorous evidence presents a significant future research demand. Those with MCI and mild AD might not respond identically to interventions with more robust evidence in the general population or in those with severe AD dementia. Sleep disturbance may well play a role in triggering or facilitating pathophysiology in AD and is the subject of current, ongoing research. Finding evidence-based treatments for sleep disturbance in a population before more severe manifestations of AD dementia develop, may therefore even offer the potential to delay the onset of more significant functional impairment.

## CONCLUSION

In the specific population of MCI and mild AD there is a relative paucity of evidence available in supporting efficacious interventions for sleep disturbance, nonetheless positive outcomes have been reported. Psychotherapeutic approaches utilising adapted CBT, melatonin and CPAP for OSA all hold promise. In our opinion, given the wide health risks of OSA, screening should become routine in memory clinics. Despite behavioural interventions often being low cost and non-toxic, we found remarkably little work in this domain. This review identifies a significant need to explore multiple, alternative sleep interventions through high quality, comparison experimental studies utilising validated sleep outcome measures.

## Data Availability

Search results for this systematic review are available on request.

https://www.crd.york.ac.uk/prospero/display_record.php?ID=CRD42019126320

## ACKNOWLEDGEMENTS

This research was funded by North Bristol NHS Trust Research Capability Funding and through contributions from Bristol Research into Alzheimer’s and Care of the Elderly (BRACE), Bristol.

With additional thanks to the North Bristol NHS Trust Library

